# A significant increase in Tuberculosis diagnosis is required to mitigate the impact of COVID-19 on its future burden

**DOI:** 10.1101/2021.07.24.21261074

**Authors:** Mario Tovar, Alberto Aleta, Joaquín Sanz, Yamir Moreno

**Affiliations:** Institute for Biocomputation and Physics of Complex Systems (BIFI), University of Zaragoza, Zaragoza 50009, Spain; Department of Theoretical Physics, University of Zaragoza, Zaragoza 50009, Spain; ISI Foundation, Via Chisola 5, 10126 Torino, Italy

## Abstract

The ongoing COVID-19 pandemic has greatly disrupted our everyday life, forcing the adoption of non-pharmaceutical interventions in many countries worldwide and putting public health services and healthcare systems worldwide under stress. These circumstances are leading to unintended effects such as the increase in the burden of other diseases. Here, using a data-driven epidemiological model for Tuberculosis (TB) spreading, we describe the expected rise in TB incidence and mortality that can be attributable to the impact of COVID-19 on TB surveillance and treatment in four high-burden countries. Our calculations show that the reduction in diagnosis of new TB cases due to the COVID-19 pandemic could result in 824250 (CI 702416-940873) excess deaths in India, 288064 (CI 245932-343311) in Indonesia, 145872 (CI 120734-171542) in Pakistan, and 37603 (CI 27852-52411) in Kenya. Furthermore, we show that it is possible to revert such unflattering TB burden scenarios by increasing the pre-covid diagnosis capabilities at least a 75% during 2 to 4 years. This would prevent almost all TB-related excess mortality caused by the COVID-19 pandemic, which will be observed if nothing is done to prevent it. Our work therefore provides guidelines for mitigating the impact of COVID-19 on tuberculosis epidemic in the years to come.

## I. INTRODUCTION

Tuberculosis (TB) is an infectious disease caused by the bacterium *Mycobacterium tuberculosis* (*M*.*tb*.) that usually affects the lungs. It is a preventable but complex disease with a high global burden that requires early detection and long treatments. Despite the global effort to eradicate TB and recent decreases in its burden due to the implementation of strategies aimed at optimizing diagnosis and treatment [1, 2], it still remains one of the greatest threats to public health worldwide, being the deadliest single-agent persistent infectious disease nowadays. According to the 2020 Global TB Report by the World Health Organization (WHO)[3], 10 million people developed TB and nearly 1.5 million people died because of TB infection during 2019. In the last decades, the WHO has deployed a series of global strategies that have since been the backbone of the global fight against TB. In 1995, the Directed Observed Treatment Strategy (DOTS) was introduced, which significantly strengthened the capacity of national programs to diagnose and treat TB cases. Later, the Stop TB Strategy, announced in 2006, was the first of such plans at setting a TB elimination horizon, defined as a reduction of incidence levels under 1 case per million and year by 2050. A redefinition of the eradication goal took place in 2014, when the previous objective was anticipated to 2035 within the End TB Strategy.

If the elimination target set by the End TB strategy was already an ambitious goal[4], the emergence of the COVID-19 pandemic caused by the new coronavirus SARS-CoV-2 sheds significant concerns into whether these goals are still reachable. During the acute stages of the COVID-19 pandemic, economic and human resources were redirected to control and mitigate the emergency caused by the pandemic, which have led to a great reduction in the diagnosis of new cases of other diseases, as already documented for cancer, or malaria [5, 6]. Interventions such as long lockdowns and mobility restrictions have exacerbated shortages in resources otherwise destined to the care of patients suffering these, and other pathologies. Moreover, COVID-19 have greatly affected healthcare workers [7–9], thus creating additional pressure to healthcare systems.

Tuberculosis diagnosis and patient care are no exceptions. As a primary and immediate effect of COVID-19 spreading onto TB transmission dynamics, a reduction in the case notification ratio has been observed during and immediately after lockdowns and periods of high COVID-19 incidence and saturation of healthcare facilities [3]. We hypotyhesize that this disruption alone will lead to a surge of TB burden in the next years, even before more complex, and less predictable effects of COVID-19 pandemic on TB management and transmission dynamics can be properly characterized. For example, according to [10–12], drastic drops in laboratory capacity needed to support TB diagnosis are expected along with interruptions in the continued supply of drugs, which could result in shortages of medications and to delaying the start of treatments until the supply chain is reestablished. Moreover, as suggested in [13], even temporary stoppages can cause long-term increases in TB incidence and mortality, and that a peak in TB burden is to be observed as a consequence of the healthcare system disruption.

In this work, we assess the impact of COVID-19 on expected TB burden until the year 2035, which marks the target horizon of the End TB Strategy. Specifically, we incorporate the observed drop on TB diagnosis and treatment compliance rates caused by COVID-19 into a mathematical model that produces long-term forecasts of TB burden [14]. This allows us to: i) quantify the effect of the COVID-19 stoppage in relation to a baseline scenario in which no pandemic happened, and ii) compute the effect that a rapid response to the uprising TB burden in the following years, in the form of a compensatory intervention aiming at boosting TB diagnosis rates as soon as the COVID-19 pandemic ends, has over long-term TB goals. Our results show that an effort focused on increasing TB diagnosis capabilities once the pandemic is over could revert the effect of the pandemic in the long term.

## II. RESULTS

### A. Forecasts of TB Incidence and mortality under Covid-19 pressure

To forecast the effect that disruptions in the diagnosis capabilities and the treatment completion have over TB incidence and mortality trends, we selected four different high-burden countries, 3 in Asia (India, Indonesia, Pakistan) and 1 in Africa (Kenya). Then, we calibrated the mathematical model described in [14] using the current WHO estimates for TB incidence and mortality rates in those countries, and produced forecasts in two separate scenarios. The baseline scenario assumes no disruption, whereas the perturbed scenario incorporates the effects of the pandemic on TB transmission dynamics. The latter effects are modeled as country-specific drops in diagnosis rates together with a reduction in TB treatment completion rates, all active for a period of 2 years. The duration of the disruptions caused by the pandemic has been set to this value because, even if the more restrictive measures (e.g. lockdowns) have been in place for briefer time-spans, disturbances over the diagnosis rate and treatment compliance are still ongoing 1.5 years after the pandemic unfolded.

Diagnosis disruption is introduced using a piece-wise function as described in Figure 1. It makes use of the calibrated parameter *d*(*t*) (see Methods, equation 3), that, as shown in equation 1, captures the diagnosis rate evolution that was expected, according to our mathematical model, prior to the irruption of COVID-19 for the period under analysis. Instead, the actual diagnosis rate in the COVID-19 scenario, denoted as *D*(*t*), considers 3 temporal intervals: up to 2020, it equals the calibrated diagnosis *d*(*t*). Then, the pandemic disruption takes place and its value is reduced to a fraction (*d*_red_) of its initial value, accordingly to reported drops for each country until the start of 2022. Afterwards, it rises back again, to values comparable to the calibrated diagnosis until the end of the simulation (2035).

**FIG. 1.**
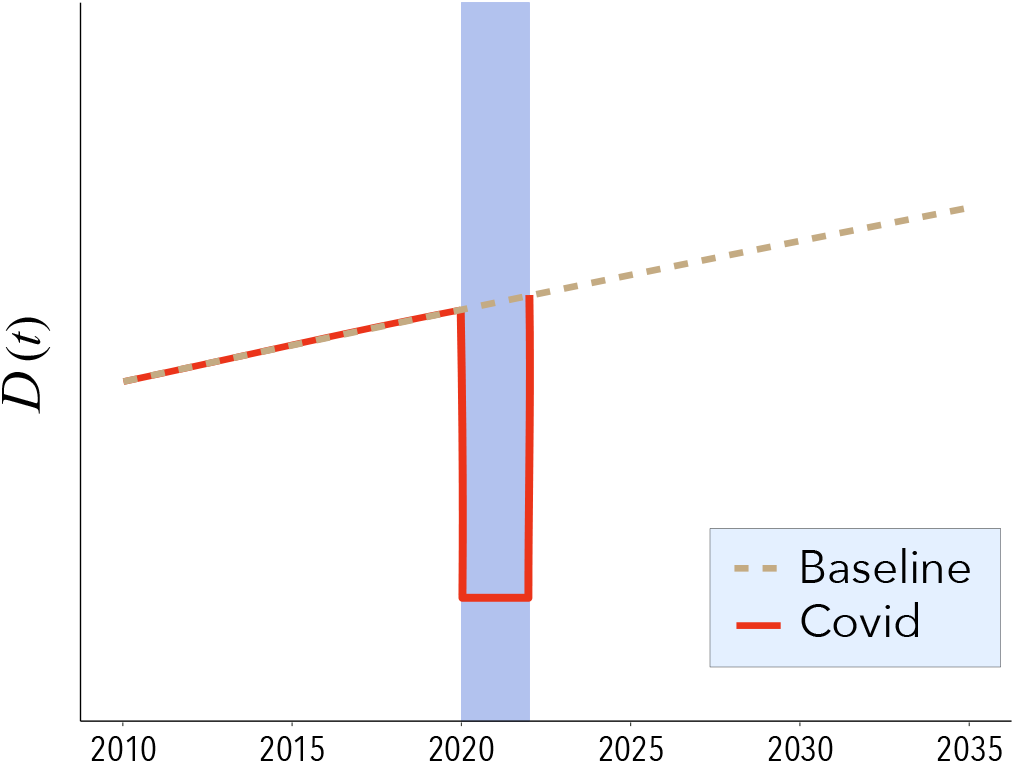
Scheme of the changes on the diagnosis rate before, during and after the pandemic period. We consider two scenarios: in the baseline, no disruption (dotted lines) are considered. In the pandemic scenario, we assume a drop characterized by a piecewise function of the diagnosis rate (salmon-shaded area) followed by a return of the diagnosis rates to the baseline scenario.

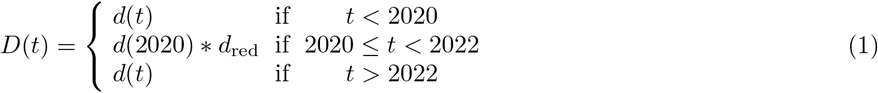

An important feature of our modeling approach is that, as shown in Eq. 1, when the pandemic regime starts, the diagnosis rate drops to a fixed value during the entire time window in which the COVID-19 disruptions are active. This modeling assumption is motivated by the severe, and country-specific decays observed in high-burden countries world-wide for TB notifications reported during the first half of 2020 (see Methods, table II). It is worth mentioning that we modeled the decrease in this way because, after calibration, in normal conditions *d*(*t*) would be an increasing function of *t* in every country under study. Therefore, applying a multiplier to *d*(*t*) directly would lead to an increase in the diagnosis rate even during lockdowns, which is clearly unrealistic as healthcare systems are not expected to improve their capabilities for TB diagnosis while devoting all efforts to control and mitigate the COVID-19 emergency. Figure 2 shows the estimated TB incidence per million inhabitants per year in the four countries considered, both in the baseline scenario and considering the negative impact of the COVID-19 pandemic. As observed, a high surge of TB incidence starts in 2020, which is followed by a pronounced peak. In the figure, the dotted line represents the baseline scenario, namely, what would have been the projected evolution of TB incidence without the disruptions of the pandemic. The size of the peak reflects the severity of the saturation of the healthcare system in each country that led to drops in diagnosis and treatment completion. The results show that the estimated COVID-19 impact on TB incidence trends is larger in the three asian countries analyzed than in Kenya. This is a direct consequence of the less severe decays in TB case notifications that have been observed in Africa with respect to other regions [3], which have been used to inform our mathematical model. These regional differences, in turn, may be due to a combination of factors. First, as stated in [15], some of those countries adopted early on measures for facing the pandemic, and secondly, COVID-19 has had a smaller effect in Africa, partially because its younger population.

**FIG. 2.**
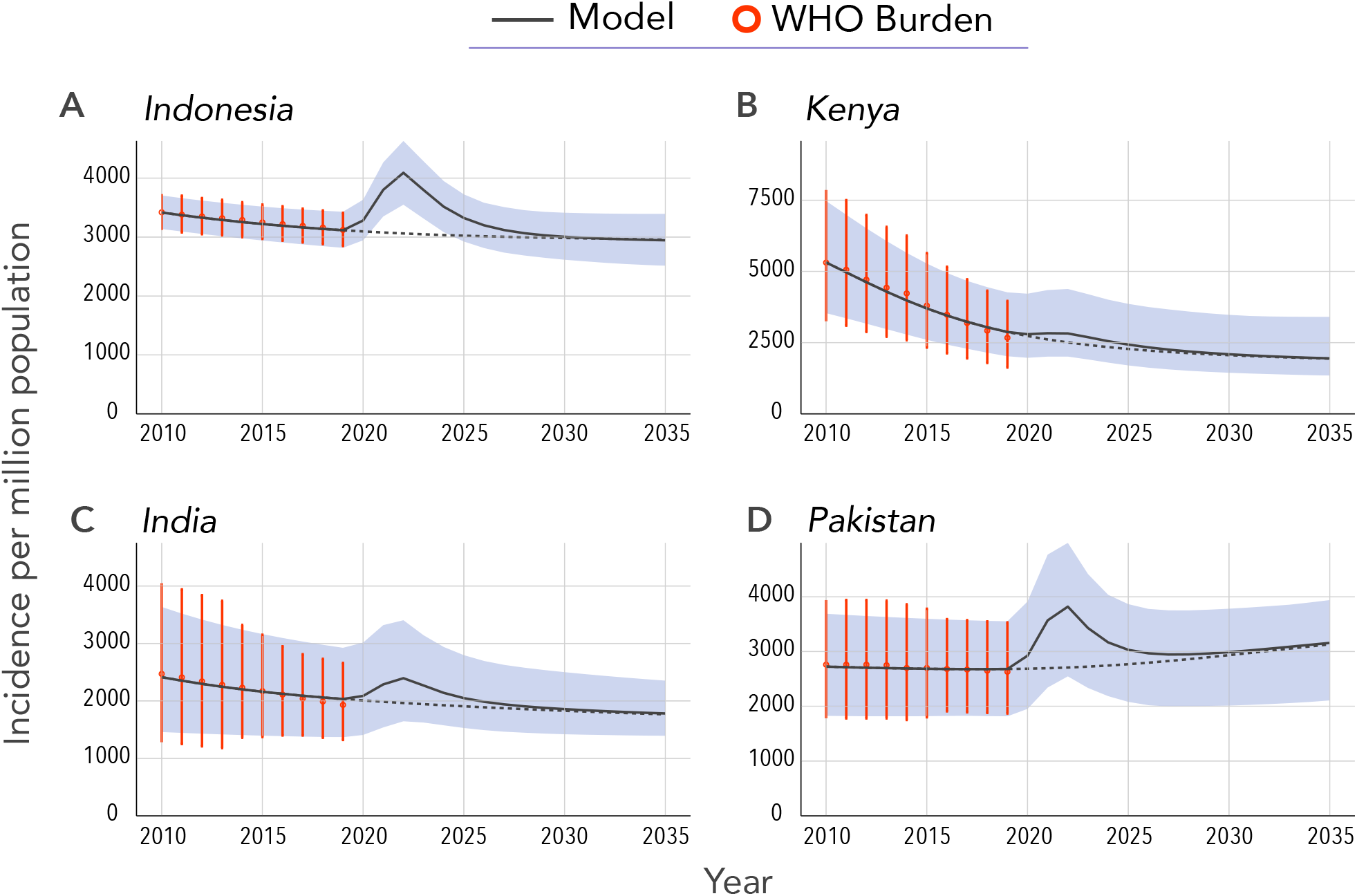
Projected annual TB incidence in four high-burden countries over the period 2020-2035. The data-driven model is calibrated with WHO incidence data up to 2019[3]. The shaded area represents the 95%CI and the black line is the median of the model outcome for disrupted scenario. The dotted black line is the model forecast for the scenario in which there was no Covid-19 pandemic. Red dots with error bars are the TB burden provided by the WHO[3] used for calibration. Projected incidence values are calculated at the end of the corresponding year in the x-axis. The impact of COVID-19 is modeled as a reduction in diagnosis rates and in treatment completion over a period of two years (2020 and 2021), see Fig. 1 and main text. The four countries considered are **A:** Indonesia. **B:** Kenya. **C:** India. **D:** Pakistan, which accounts for 42.1% of the total number of TB infections worldwide.

Important enough, even if COVID-19 disruptions are assumed to last only 2 years, the long-term effects span for longer times, a decade or more since the start of the COVID-19 pandemic. As observed in figure 2, in the long term, TB incidence levels stabilize and recover to their baseline values approximately by the year 2030, resulting in a 10 years window of higher burden that makes the incidence to go off the way of TB eradication stated in the End TB Strategy. Moreover, in the absence of any further intervention, the peak of TB incidence caused by the disruptions associated to the COVID-19 pandemic will produce not only new TB cases but also an increase in TB related deaths all across the world. Specifically, by the end of the simulation period in the year 2035, our model predicts an increase in mortality as shown in figure 3, where we have represented both the increment percentage and the total number of accumulated additional deaths between 2020 and 2035. Particularly, our forecast predicts an increase in the number of deaths of 4.65%(3.96-5.55, 95%CI) in India, 7.64%(6.58-9.05, 95%CI) in Indonesia, 2.09%(1.91-2.36, 95%CI) in Kenya and 8.68%(7.13-10.17, 95%CI) in Pakistan. In absolute terms, the total number of excess deaths could be over 1.29 million individuals in these four countries alone (Figure 3B).

**FIG. 3.**
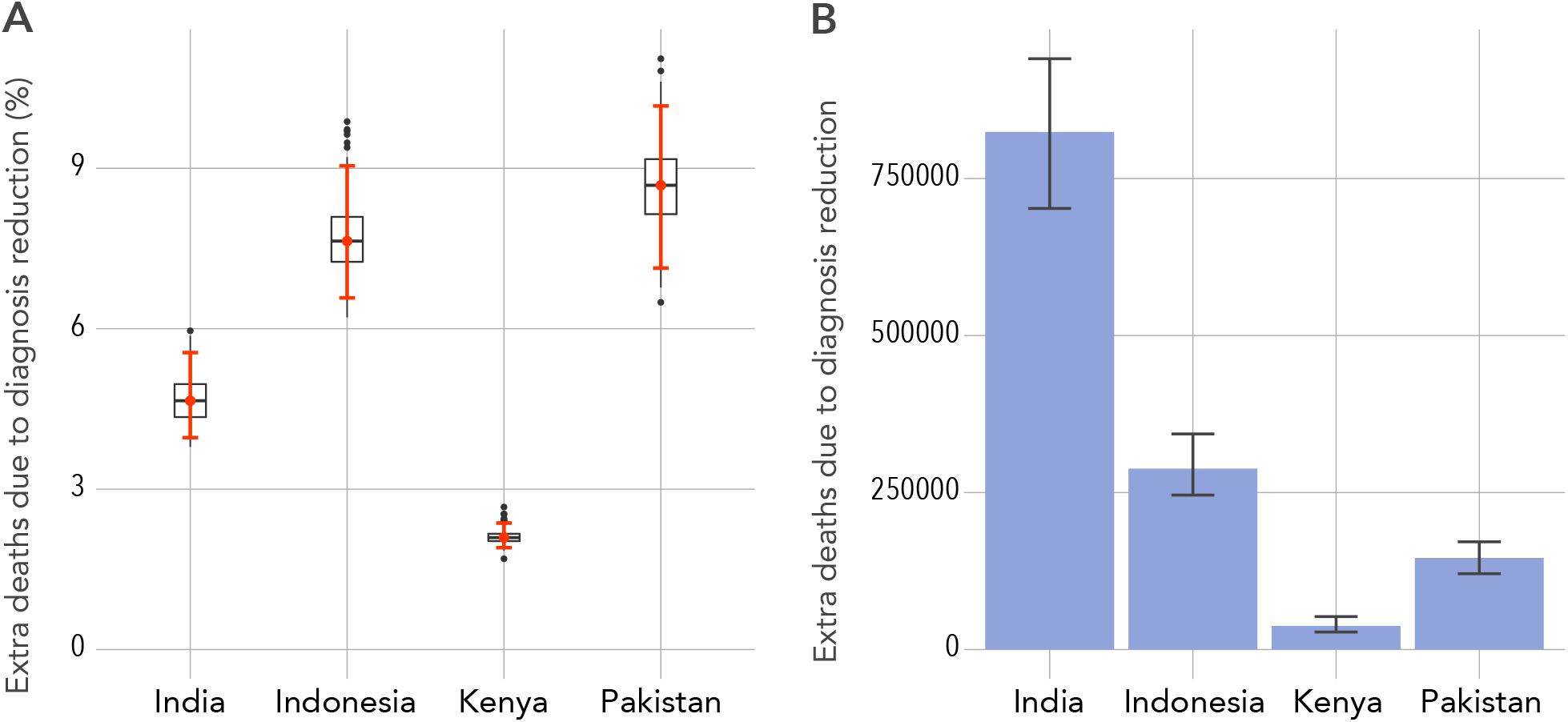
Model predictions of additional TB related deaths due to COVID-19 related disruptions in healthcare systems. Error bars are the 95%CI. **A:** Percentage of increase of mortality with respect to the baseline scenario in 2035 for each of the four countries studied. **B:** Cumulative number of excess deaths cause by the pandemic impact during the whole time window simulated (2020-2035) for each country under study as indicated in the x-axis.

### B. Proposed intervention for mitigating the pandemic effect

As shown before, the ongoing pandemic situation will lead to significant increases in TB incidence and mortality. This represents a critical setback with respect to the mid-to-long-term objective of eradicating TB disease within the next few decades, making it hardly achievable without a rapid and effective recovery strategy. More importantly, the disruptions will cause many preventable deaths. It is thus of utmost importance to elucidate whether new policies could be implemented to revert the negative impact of COVID-19 in TB disease. In what follows, we show that it is indeed possible to shorten the estimated decade that would be required to regress to pre-pandemic TB projections, avoiding almost all excess deaths by actively boosting TB diagnosis rates, at least temporarily, to levels higher than those observed before COVID-19. Additionally, given that we assume that during this phase the healthcare systems are not saturated any more, pre-pandemic levels for treatment compliance are also recovered once the COVID-19 disruptions are switched off.

The potential intervention over the diagnosis of TB cases is modeled using the same piece-wise function of Eq. 1 but with an additional piece introduced after the pandemic disruption is over and for a parameterized duration, T_rec_ − 2022, to be determined. More specifically, we assume that over this new period of time the pre-pandemic diagnosis rate is effectively increased by a factor *d*_inc_ *≥* 1, (see also figure 4) that is:

**FIG. 4.**
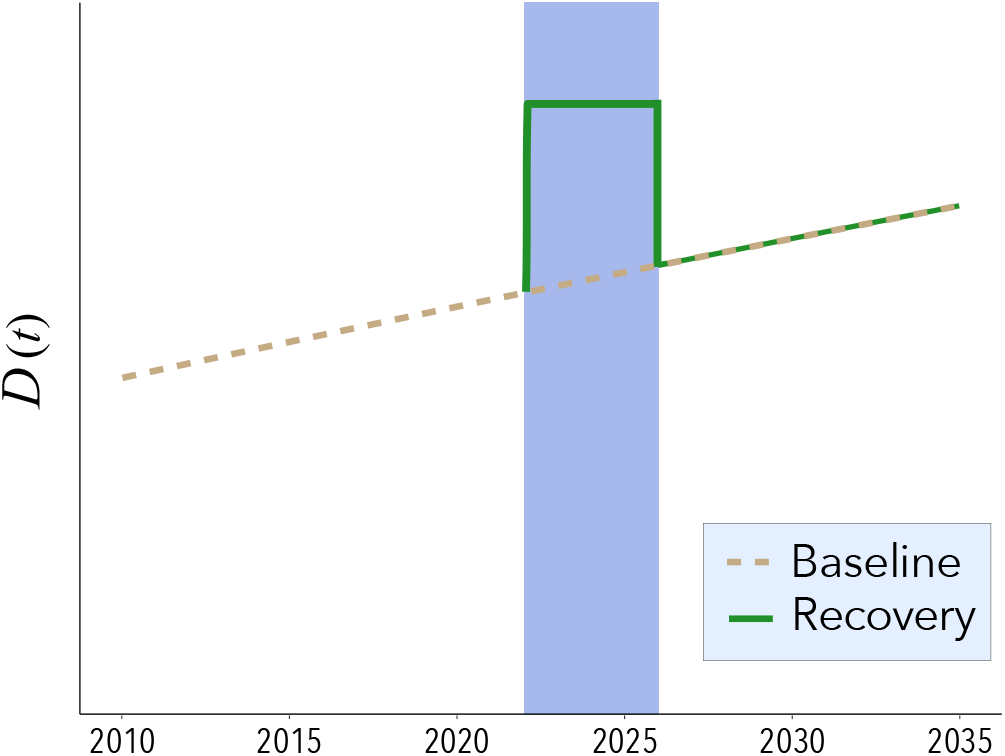
Schematic representation of the increase in diagnosis rate in the post-pandemic scenario. Unlike the situation described in figure 1, we consider a compensatory period of duration between 2022 and T_*rec*_ during which the diagnosis rate is boosted up to *d*(2020) * *d*_*inc*_, with *d*_*inc*_ *>* 0.

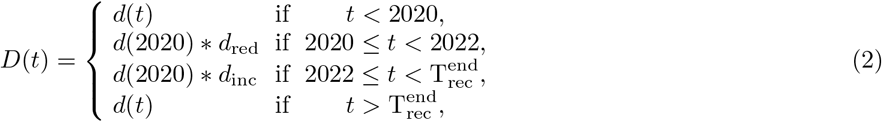

A scheme of the evolution of the diagnosis rate under this alternative scenario is shown in figure 4 (mauve-shaded area).

The efficacy of the introduction of such compensatory period will in principle be proportional to the intensity and the duration of the intervention, namely *d*_inc_ ≥ 1 and 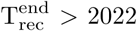. In figure 5, we show the impact of considering different combinations of these two parameters on the cumulative excess mortality in 2035. Clearly, the more intense and longer the additional effort is, the larger the number of averted deaths by the end of the simulation period. As it can be seen, except for Pakistan, for the rest of the countries, an increase in diagnosis rate during a certain amount of time could eventually revert the negative impact of COVID-19 in TB mortality measured in 2035. More specifically, for Indonesia, Kenya and India, there is a region in the parameter space for which the increase in mortality in 2035 is close to zero (upper-right corners of the different panels) if the diagnosis rate of new TB cases nearly doubles for a period that spans between 2 and 4 years. Importantly, our simulations also show that no matter the increase in diagnosis rate and its duration, not all countries would be able to avoid the impact of COVID-19 disruption on TB mortality. Admittedly, our results estimate that by 2035, the excess number of TB-related deaths caused by the pandemic in Pakistan will be at least 2%, even for a best-case scenario that assumes a long and very intense recovery strategy. For such countries, additional – and different in nature– interventions are needed. On the positive side, we also observe in panel B of Fig. 5 that for countries like Kenya, it is not only possible to evade the impact of the pandemic, but to reduce the expected number of TB-related deaths by 2035 below the baseline (i.e., no pandemic) scenario, at least considering the information in our hands as of today.

**FIG. 5.**
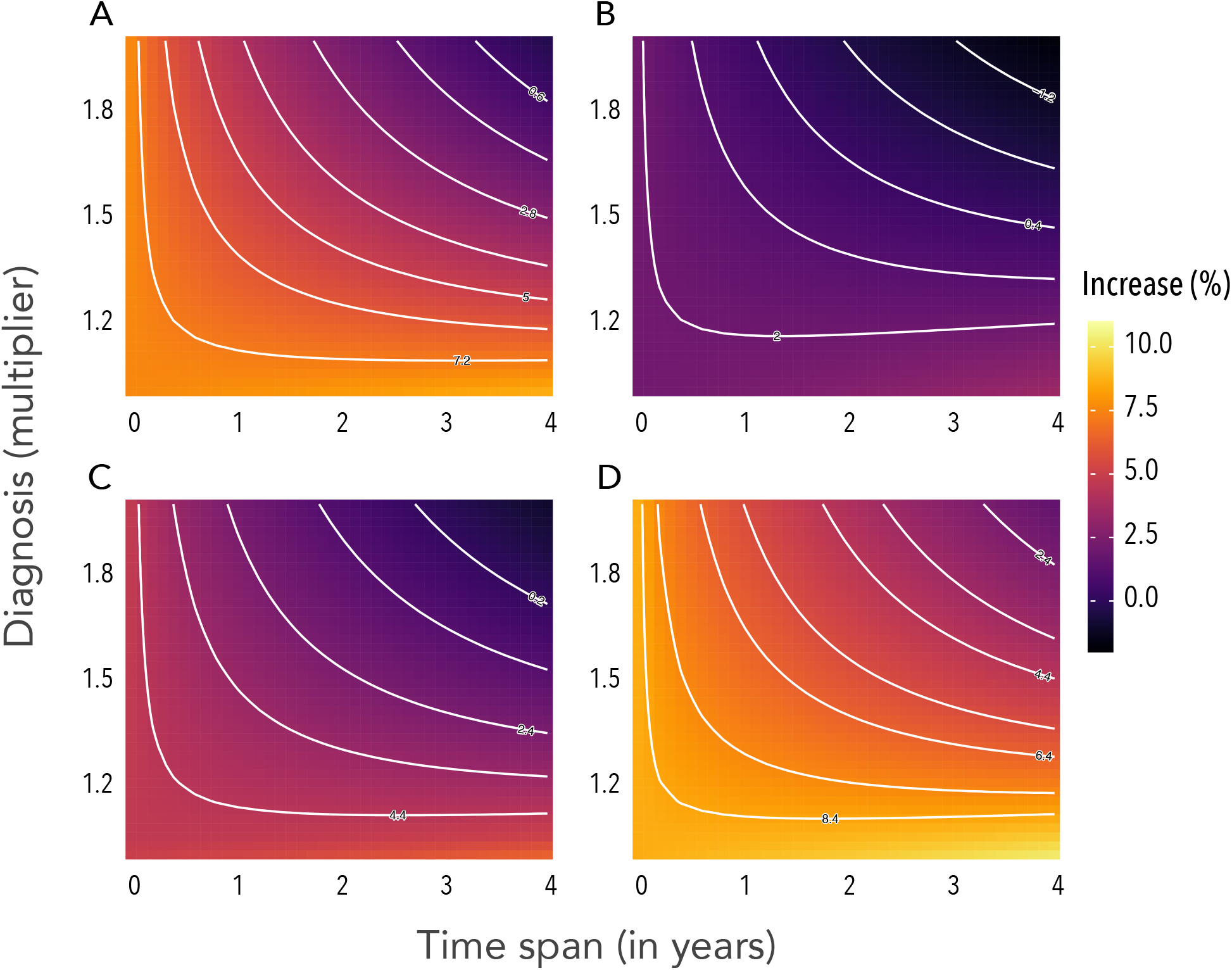
Relative increase of the expected number of deaths when an intervention is introduced in 2022 with respect to the projected impact of the COVID-19 pandemic. We assume that during the recovery period, whose duration is given by the parameter T_rec_ (the values of the X axis), there is an increase in the diagnosis rate characterized by a factor *d*_inc_ *>* 1 (value of the Y axis). Level (white) curves represent combinations of parameters that give the same excess deaths percentage, as indicate by the values over the curves. The four countries under study are **A:** Indonesia. **B:** Kenya. **C:** India. **D:** Pakistan.

**FIG. 6.**
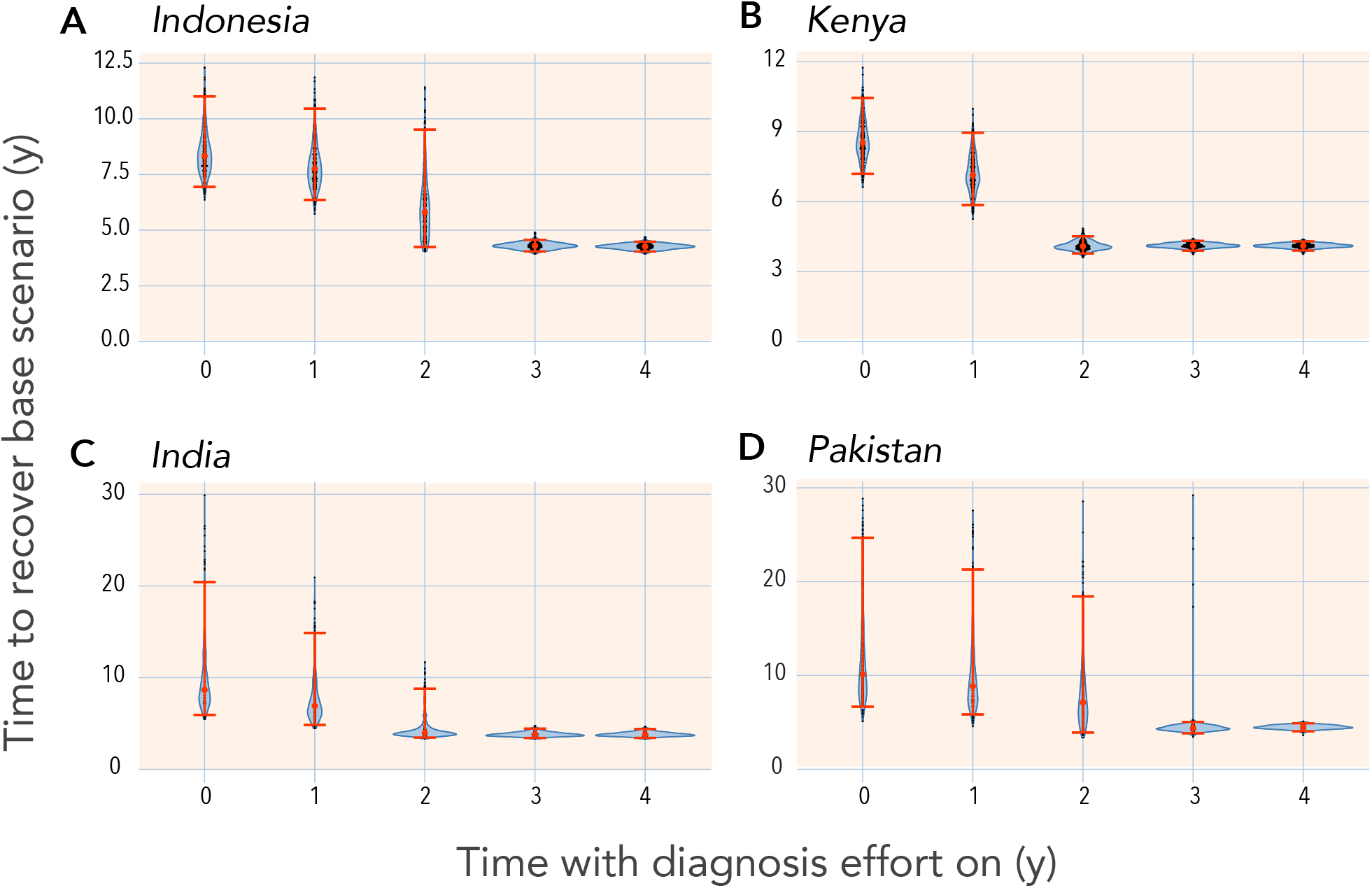
Time to recover the baseline scenario (indicated in the y-axis) under different durations (given in the x-axis) of the non-pharmaceutical intervention proposed described in this study. Results shown correspond to an increase in diagnosis of 50%. The time it takes for the incidence to regain the levels with no pandemic is calculated as the point at which projected incidence curves for baseline and disrupted scenarios cross with each other. Error bars are the 95%CI. The four countries studied and shown in the different panels are **A:** Indonesia. **B:** Kenya. **C:** India. **D:** Pakistan.

Table I reports the number of averted deaths in each country in 2035 when the additional effort is applied during two to four years and considering increases in the diagnosis rates of 25%, 50% and 75%, starting in 2022. The reported values are obtained comparing model forecasts for the estimated number of TB-related deaths in the pandemic scenario with the outcome obtained when the recovery strategy is put in place after the end of the COVID-19 disruptions. The model suggests that it is generally better to increase more the diagnosis rate for shorter times than increasing the temporal span and having smaller increments of the diagnosis rate, because in the former situation, more deaths are averted in the long term. Nonetheless, the ideal scenario is still the one in which both dimensions are boosted at the same time, as the longer the time the effort is maintained for a given multiplier, the lower the TB-related death toll caused by the pandemic. As noted before, we stress that for an effort ratio of 1.75 and a temporal span of 4 years, the number of averted deaths almost corresponds to the 100% of additional deaths expected due to the COVID-19 disruptions (see figure 3), i.e., the pandemic impact could be fully mitigated.

**TABLE 1.** Cumulative number of averted deaths in 2035 with a post COVID-19 intervention initiated in 2022 in each of the countries studied. The values in the table are computed by calculating the difference between the model forecast for mortality with the pandemic scenario and with non-pharmaceutical interventions of different intensities of diagnosis effort and duration of the recovery period. Values are the median of the outcome and figures in parentheses are the 95% CI of the model projections.

| Country | Diagnosis effort | T2y | T3y | T4y |
| --- | --- | --- | --- | --- |
| Indonesia | 1.25 | 59932(51052-71961) | 77701(66342-93055) | 92272(78972-110083) |
|  | 1.50 | 117010(98977-140541) | 151591(128685-181185) | 181625(155086-215409) |
|  | 1.75 | 160674(136251-192328) | 206357(175931-245357) | 246385(211468-290944) |
| Kenya | 1.25 | 7287(5347-10715) | 8296(6084-12351) | 8117(5936-12254) |
|  | 1.50 | 21839(16070-30851) | 28258(20702-40029) | 33048(24124-47092) |
|  | 1.75 | 34176(25119-47826) | 44529(32581-62405) | 52828(38488-74375) |
| India | 1.25 | 172233(144920-203580) | 219970(185718-259857) | 255907(216146-301548) |
|  | 1.50 | 363277(295658-455633) | 472507(386375-595902) | 565112(466075-709222) |
|  | 1.75 | 511338(408676-669979) | 660454(533543-873433) | 790709(647066-1038902) |
| Pakistan | 1.25 | 22133(18137-26343) | 27813(22708-33092) | 31818(25920-38259) |
|  | 1.50 | 46003(37857-53383) | 59202(49365-68666) | 70523(59432-81865) |
|  | 1.75 | 63360(52093-73029) | 81507(68110-93937) | 97800(82825-112126) |

### C. Time to recover baseline scenario

As the previous calculations show, implementing a strategy aiming at boosting temporarily TB diagnosis rates during a compensatory period constitutes a solution potentially able to revert the negative impact of the COVID-19 pandemic on TB excess mortality. However, it is also key to reduce TB incidence levels. To shed light on the effectiveness of such intervention on reducing TB incidence, we evaluate how much time it would take to recover the baseline, pre-pandemic projections for TB incidence with the intervention proposed, as a function of the duration of the compensatory period. Results for several scenarios and the four countries under study are shown in figure First, we explore the case in which nothing is done after the pandemic is over, which provides a reference for the no-intervention scenario. For the rest of scenarios, we assume that a compensatory period is in place with an effective increase in diagnosis rate of 50% and we explore how the temporal span of the effort modifies future burden projections.

Model forecasts suggest that a temporal span of 2 to 4 years with a 50% boost on diagnosis is enough to restore the annual incidence level of the baseline scenario in around 5 years, for all countries. Note that even for larger recovery periods, the time needed to regain the basal incidence could not be lowered further, since boost in the the diagnosis rate needs to be larger if that goal is to be pursued. All together, our results show that a relative large and intense recovery strategy focused on increasing diagnosis rates above pre-pandemic standards during a short period of time would permit the reversion of most of COVID-19 impact on TB incidence and mortality rates alike.

## III. DISCUSSION

The COVID-19 pandemic is not over yet, and much remains to be clarified about its impact over the −physical and mental − health of the general population. As of July, 2021, the novel coronavirus SARS-CoV-2 has infected more than 185 million individuals, causing the death of more than 4 million persons worldwide. Being this a novel coronavirus and disease, the scientific community has already been able to describe many of the clinical characteristics and pathogenesis of COVID-19, specially during the acute phase [16, 17]. However, there are important features that remain less known, such as the long-term consequences of the disease [18–20] and the relation between comorbidities and their risks upon infection by SARS-CoV-2 [21, 22]. Another important question that is not fully assessed concerns the indirect effects of the pandemic, and the NPIs adopted for its control and mitigation, over other diseases.

In particular, the large number of healthy individuals that were infected in a very short period of time, producing the so-called epidemic waves, led to the saturation of many healthcare systems, which in turn induced the implementation of very restrictive measures such as lockdowns and curfews in those countries. These compulsory interventions have been argued to be at the root of important reductions in diagnosis rates of other deadly diseases, as shown in [5, 6]. Yet, the long-term consequences are still to be determined. Here, we have focused on Tuberculosis, since it is one disease for which disruptions in health care could be most dramatic [11, 13] given that even without a pandemic scenario, more than 1.5 million lives are lost every year because of the disease.

Using a data-driven epidemiological model [14], we have quantified the negative impact of COVID-19 on TB diagnosis and its long-term consequences. We have also shown that a rapid and intense intervention could eventually mitigate the expected increase in incidence and mortality of tuberculosis. Countries enrolled in this work have been selected because of their high TB burden, contributing an important amount of cases to the annual TB incidence recorded by the WHO global report. Certainly, all four countries together accounted for a 42.1% of the global TB cases in the year 2019. Individually, India comprises the 26.5%, Indonesia accounts for the 8.47%, Kenya represents the 1.40% and Pakistan is responsible for a 5.71% of all cases globally. Additionally, for these countries, the reduction in TB case notification due to the COVID-19 pandemic has been documented [3], and span over milder (Kenya) to more severe magnitudes (Indonesia), which make them suitable case studies to estimate the pandemic negative impact and the design of corrective interventions.

Our results show that a drop in diagnosis rates and in first-line treatment compliance statistics lead to a pronounced increase of TB incidence with respect to the baseline scenario. This surge could span a decade if nothing is done, even when considering that disruptions last only 2 years. In turn, the growth in TB burden leads to an upsurge in mortality, producing more than 1.29 million excess deaths by 2035 in the four countries of the study combined. However, our study also shows that most of these deaths can still be prevented. In particular, our projections show that an increase of available diagnosis capabilities for some time has very positive effects on long-term TB burden because the pandemic effect can be greatly curtailed and almost a 100% of the expected excess deaths can be avoided. It is worth stressing that the intervention proposed here is aimed at increasing the volume of diagnosed individuals, thus bringing them to treatment as soon as possible. This ultimately points towards cutting TB transmission to a point wherein pre-pandemic burden levels are recovered. As we have demonstrated here, one such intervention could be enough for full mitigation of the negative impact of COVID-19 pandemic on TB incidence and mortality.

In closing, we also mention that our approach is not exempt from limitations that affect TB transmission models. For instance, the outcome of our model depends on a series of epidemiological parameters and initial burden estimates that are subject to strong sources of uncertainty, thus propagating this uncertainty to the results. This means that future improvements in measuring the input data are expected to impact the quantitative outcomes of our mathematical model, the same way as it would affect any other model who leans on them. Moreover, in our work we have only described the disruption caused by the COVID-19 pandemic on the TB care system via a reduction of diagnosis capabilities and treatment availability. Even if these are arguably the primary, and have been the first effects of COVID-19 pandemic on TB transmission dynamics in being characterized, there may be many other effects that are yet hard to parameterize. On the one hand, it is well known that the emerging pandemic has disrupted profoundly the age-structure of social contacts in human populations world-wide through a combination of mobility restrictions, lockdowns, social distancing, and adaptive conducts driven by self-perceived risk, often associated to the stark variations in susceptibility to severe disease and death that have been extensively reported for COVID-19. All these effects combined have arguably re-wired age-dependent contact structures in a way that is not fully understood, and may not be completely transient. On the other hand, geo-political and economic shifts driven by the pandemic will for sure exert differential effects on TB transmission dynamics between countries. While the TB modeling community should commit to characterize these phenomena in depth and incorporate them into model forecasts, these are all questions that remain beyond the scope of this study. Be it as it may, the model projections reported here point towards a worrying scenario about the effects of the current pandemic on TB burden evolution in the near future, regardless of the detailed implementation of the disruptions. As more data on the possibly disparate effects of COVID-19 pandemic on TB is reported [3], updated modeling scenarios can be considered. Similarly, while the duration of the pandemic has been selected to be 2 years for all countries under study, longer estimates of this parameter could lead to an increase in the quantitative outcomes reported here.

In summary, our work shows that implementing a strategy aiming at boosting TB diagnosis rates during a compensatory period after the pandemic holds the promise of mitigating, if not fully reverting, the negative impact of the COVID-19 pandemic on TB excess incidence and mortality, even if that period of boosted diagnosis is transient. While the importance of early diagnosis to arrest TB transmission is well known in TB epidemiology [23, 24], we describe here how pushing that aspect of global TB management strategies in the early post-covid time has the potential of reverting a large fraction of the negative impact caused by the pandemic on the global TB-epidemics. Interventions such as chronic cough screenings among people seeking healthcare, or even active screening of TB cases among non-symptomatic individuals, along with protocols targeting specifically pre-clinical and/or smear-negative TB cases do all hold the potential of boosting early diagnosis rates in a way that may well be compatible with the scenarios modelled in this study [23–26]. In order to prevent the COVID-19 pandemic from destroying all the progress achieved during the last years in global TB control, it is time to prioritize such interventions.

## IV. MATERIALS AND METHODS

### A. Model calibration and diagnosis rate

In this study, we have capitalized on the detailed *M*.*tb*. transmission model described in [14]. Conceptually, this model describes TB dynamics within a whole, closed population, stratified into 15 age groups during time spans of the order of several decades. The model is detailed enough as to include demographic evolution and ageing, along with heterogeneous contact patterns among age groups that have been adapted from empirical survey studies as extensively described in [14].

Here, the model is calibrated to reproduce TB incidence and mortality rates in each country under study for the period 2000–2019, using the burden estimates provided by the WHO. The calibration process gives the diagnosis rate *d*(*t*) and the scaled infectiousness *β*(*t*), which are modeled as half-sigmoid-like curves, which, among other parameters, are country specific. This allows the model to reproduce different epidemiological scenarios. Specifically, the diagnosis rate is defined in equation 3:

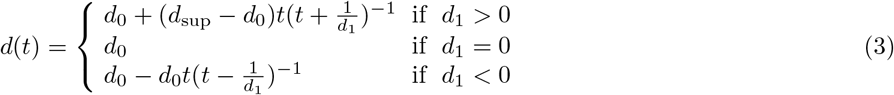

Therefore, the diagnosis rate is parameterized by two quantities (*d*_0_, *d*_1_), where *d*_0_ is the value at the beginning of the calibrating window (i.e. year 2000 in this study), and *d*_1_ defines its evolution, either increasing or decreasing with time depending on *d*_1_’s sign. In the case of a decreasing evolution, the diagnosis rate is bounded to be greater than zero, while in the case of increasing evolution the upper bound is *d*_sup_ = 12.17y^−1^[14]. This latter upper bound corresponds to a minimum diagnosis period of one month, assuming that, with a conservative lower boundary, the main symptom of TB is a continuous cough lasting for three weeks, followed by a time to diagnose estimated to last at least 10 days [27]. For further details regarding the specific values of epidemiological parameters, calibration processes and uncertainty estimates, the reader is referred to [14].

Once the model is calibrated, we use it to produce forecasts until 2035 under two different scenarios: the baseline scenario, namely, an scenario in which there is no COVID-19 pandemic and thus, no disruption in healthcare systems is introduced, and another one in which a disruption is introduced at the start of 2020 up to the end of 2021, which is the pandemic scenario. During the duration of the pandemic, the diagnosis rate drops accordingly to the reduction observed in the notifications of TB cases in the first half of 2020, provided in the last global TB report, thus, being country specific, see Table II. Finally, during the period of recovery, interventions are aimed at compensating the drop in diagnosis rates of the pandemic years. We modeled this by multiplying the diagnosis by a scale parameter, as already discussed. Once the recovery period is over, we assume that the diagnosis rate goes back to its original value as given by *d*(*t*) up to the end of the simulation.

**TABLE II.** Multiplier for the diagnosis reduction in selected countries. The value of the multiplier is extracted from the global TB report (2020) using the TB notification drop data in each country.

| Country | Diagnosis reduction multiplier ($d_{\text{red}}$) | Source |
| --- | --- | --- |
| India | 0.40 | WHO (2020) |
| Indonesia | 0.30 | WHO (2020) |
| Kenya | 0.60 | WHO (2020) |
| Pakistan | 0.35 | WHO (2020) |

### B. First-line treatment reduction

According to [3, 13], first-line TB treatment completion has dropped effectively as a consequence of the COVID-19 pandemic, with interruptions in the supply of drugs that delay the start of the treatment in those cases in which the remaining medical capabilities have been enough to diagnose the disease. This inconvenience could not only worsen the expected treatment outcome for the patient, but also drive secondary infections even in diagnosed patients if they are not able to quarantine until the treatment could be carried out. We modeled this situation in terms of the epidemiological model by including a fraction of under-treatment pulmonary TB individuals (*T*_*p*_) in the expression of the force of infection (*λ*(*t*)). On the baseline scenario and without disruptions, those *T*_*p*_ individuals are not able to contribute to *λ*(*t*) as we assume them to be under control by the healthcare system, thus, being controlled and either under quarantine or, later on, medicated with TB drugs that greatly reduce their infectiousness. This means that, under normal circumstances, diagnosed individuals are expected not to be a risk for the rest of the population. However, when disruptions on the supply chain appear, a drop is observed in the first-line and second-line treatments completion [13], and then diagnosed individuals who are not able to either start the treatment or quarantine could become a risk. For this reason, we obtain an estimate of the fraction of *T*_*p*_ individuals that contribute to *λ*(*t*), *T*_inf_, from Cilloni et al. [13] as

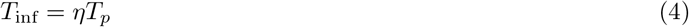

where *η* = 0.788. This value attempts to capture this kind of impact in countries like India and Kenya. It is based on expert opinion in the Stop TB Partnership and USAID about the side effects of the COVID-19 pandemic on TB treatment completion. We assume it to be a good proxy for the real value for the other countries included in this study.

## Data Availability

Data used in the study are publicly available in WHO repositories.

## Acknowledgments

M.T. acknowledges the support of the Government of Aragón through a FPI contract. A. A and Y.M. acknowledges partial support from the Government of Aragon and FEDER funds, Spain through grant E36-20R (FENOL), and by MINECO and FEDER funds (FIS2017-87519-P). A.A. and Y.M. acknowledge support from Banco Santander (Santander-UZ 2020/0274) and the financial support of Soremartec S.A. and Soremartec Italia, Ferrero Group. J.S. acknowledges support from the Spanish Ministry of Science and Innovation (MICINN) through grant PID2019-106859GA-I00 and Ramón y Cajal research grant RYC-2017-23560. The funders had no role in study design, data collection, and analysis, decision to publish, or preparation of the manuscript.

## References

[1] M. W. Borgdorff, K. Floyd, and J. F. Broekmans, Bulletin of the World Health Organization 80, 217 (2002).

[2] A. Matteelli, A. Rendon, S. Tiberi, S. Al-Abri, C. Voniatis, A. C. C. Carvalho, R. Centis, L. D’Ambrosio, D. Visca, A. Spanevello, and G. B. Migliori, European Respiratory Review 27, 10.1183/16000617.0035-2018 (2018).

[3] W. H. Organization, Global tuberculosis report 2020 (2020).

[4] C. Lienhardt, P. Glaziou, M. Uplekar, K. Lonnroth, H. Getahun, and M. Raviglione, Nature Reviews Microbiology 10, 407 (2012).

[5] M. Richards, M. Anderson, P. Carter, B. L. Ebert, and E. Mossialos, Nature Cancer 1, 565 (2020).

[6] R. Ansumana, O. Sankoh, and A. Zumla, Nature Medicine 26, 1334 (2020).

[7] J. A. Gold, Covid-19: adverse mental health outcomes for healthcare workers (2020).

[8] Y. Chen, X. Tong, J. Wang, W. Huang, S. Yin, R. Huang, H. Yang, Y. Chen, A. Huang, Y. Liu, et al., Journal of Infection 81, 420 (2020).

[9] F. Fusco, M. Pisaturo, V. Iodice, R. Bellopede, O. Tambaro, G. Parrella, G. Di Flumeri, R. Viglietti, R. Pisapia, M. Carleo, et al., Journal of Hospital Infection 105, 596 (2020).

[10] W. R. O. for Europe, Rapid communication on the role of the gen-expert Ó sars-cov-2 in the who european region (2020).

[11] P. Adepoju, The Lancet HIV 7, e319 (2020).

[12] platform for rapid molecular testing for A.A. Malik, N. Safdar, S. Chandir, U. Khan, S. Khowaja, N. Riaz, R. Maniar, M. Jaswal, A. J. Khan, and H. Hussain, Health policy and planning 35, 1130 (2020).

[13] L. Cilloni, H. Fu, J. F. Vesga, D. Dowdy, C. Pretorius, S. Ahmedov, S. A. Nair, A. Mosneaga, E. Masini, S. Sahu, et al., EClinicalMedicine 28, 100603 (2020).

[14] S. Arregui, M. J. Iglesias, S. Samper, D. Marinova, C. Martin, J. Sanz, and Y. Moreno, Proceedings of the National Academy of Sciences 115, E3238 (2018), https://www.pnas.org/content/115/14/E3238.full.pdf.

[15] N. Haider, A. Y. Osman, A. Gadzekpo, G. O. Akipede, D. Asogun, R. Ansumana, R. J. Lessells, P. Khan, M. M. A. Hamid, D. Yeboah-Manu, et al., BMJ Global health 5, e003319 (2020).

[16] W. J. Wiersinga, A. Rhodes, A. C. Cheng, S. J. Peacock, and H. C. Prescott, Jama 324, 782 (2020).

[17] M. Cevik, K. Kuppalli, J. Kindrachuk, and M. Peiris, bmj 371 (2020).

[18] M. Marshall, Nature 594, 168 (2021).

[19] C. Huang, L. Huang, Y. Wang, X. Li, L. Ren, X. Gu, L. Kang, L. Guo, M. Liu, X. Zhou, et al., The Lancet 397, 220 (2021).

[20] C. H. Sudre, B. Murray, T. Varsavsky, M. S. Graham, R. S. Penfold, R. C. Bowyer, J. C. Pujol, K. Klaser, M. Antonelli, L. S. Canas, et al., Nature medicine 27, 626 (2021).

[21] A. Sanyaolu, C. Okorie, A. Marinkovic, R. Patidar, K. Younis, P. Desai, Z. Hosein, I. Padda, J. Mangat, and M. Altaf, SN comprehensive clinical medicine, 1 (2020).

[22] B. Wang, R. Li, Z. Lu, and Y. Huang, Aging (Albany NY) 12, 6049 (2020).

[23] P. M. Small and M. Pai, New England Journal of Medicine 363, 1070 (2010), publisher: Massachusetts Medical Society eprint: https://doi.org/10.1056/NEJMe1008496.

[24] K. Lönnroth, K. G. Castro, J. M. Chakaya, L. S. Chauhan, K. Floyd, P. Glaziou, and M. C. Raviglione, The Lancet 375, 1814 (2010).

[25] A. S. Azman, J. E. Golub, and D. W. Dowdy, BMC Medicine 12, 216 (2014).

[26] J. N. Sekandi, D. Neuhauser, K. Smyth, and C. C. Whalen, The International Journal of Tuberculosis and Lung Disease 13, 508 (2009).

[27] S. J. Millen, P. W. Uys, J. Hargrove, P. D. Van Helden, and B. G. Williams, PloS one 3, e1933 (2008).

